# Safety and Immunogenicity of SARS-CoV-2 S-2P Protein Vaccine MVC-COV1901 in People Living with HIV

**DOI:** 10.1101/2021.12.08.21267439

**Authors:** Shu-Hsing Cheng, Chia En Lien, Szu-Min Hsieh, Chien-Yu Cheng, Wang-Da Liu, Ching-Lung Lo, Wen-Chien Ko, Yen-Hsu Chen, Ching-Tai Huang, Hsiao-Ting Chang, Shinn-Jang Hwang, Ning-Chi Wang, Ming-Che Liu, Yu-Lin Lee, I-Chen Tai, Josue Antonio Garcia Estrada, Tzou-Yien Lin, Wen-Sen Lee

## Abstract

**Objectives:** To provide data on the immune response to COVID-19 vaccines in people living with HIV (PWH), MVC-COV1901, a recombinant protein vaccine containing S-2P protein adjuvanted with CpG 1018 and aluminium hydroxide, was assessed.

**Methods:** A total of 57 PWH of ≥ 20 years of age who are on stable antiretroviral therapy and with CD4^+^ T cell ≥ 350 cells/mm^3^ and HIV viral load < 10^3^ copies/ml were compared with 882 HIV-negative participants. Participants received 2 doses of MVC-COV1901 28 days apart. Safety and the immunogenicity were evaluated.

**Results:** No vaccine-related serious adverse events (SAEs) were recorded. Seroconversion rates (SCRs) of 100% and 99.8% were achieved in people living with HIV (PWH) and comparators, respectively, 28 days after second dose. The geometric mean titers (GMTs) (95% confidence interval [CI]) against wild type SARS-CoV-2 virus were 136.62 IU/mL (WHO Standardized International Unit) (95% CI 114.3-163.3) and 440.41 IU/mL (95% CI 421.3-460.4), for PWH and control groups, respectively, after adjusting for sex, age, BMI category, and comorbidity, and the adjusted GMT ratio of comparator/PWH was 3.22 (95% CI 2.6-4.1). A higher CD4/CD8 ratio was associated with a higher GMT (R=0.27, p=0.039).

**Conclusions:** MVC-COV1901 has shown robust safety but weaker immunogenicity responses in PWH. As a result, a third dose or booster doses of MVC-COV1901 may be appropriate for PWH.

## Introduction

The Coronavirus Disease 2019 (COVID-19), caused by the novel coronavirus SARS-CoV-2, was first reported in Wuhan, Hubei Province, People’s Republic of China (PRC) in late December 2019. The disease spread rapidly around the globe and was declared a pandemic by the WHO on March 11^th^, 2020.^1,2^ As of Nov. 2021, nearly 260 million infections and over 5 million deaths had occurred worldwide.^3^ Unfortunately, the disease has the most severe impact on people with poor socioeconomic status, and that includes populations vulnerable to HIV. ^3,4^

People who are living with HIV (PWH) are subject to many risk factors that predispose for severe COVID-19. These factors include high rates of smoking, drug use, comorbidities including cardiovascular diseases, diabetes mellitus, chronic renal diseases, chronic liver diseases, and lung diseases.^5-8^ Furthermore, many PWH are overweight or obese, which correlates with the widespread use of second generation integrase inhibitors ^9,10^ and is an additional risk factor for severe COVID-19.^11,12^ Health disparities due to socioeconomic status and stigmatization may also hinder early diagnosis of SARS-CoV-2 infection and timely provision of healthcare.^4^ Several case series and meta-analysis also pointed towards the association between PWH and severe COVID-19 and higher mortality rate. The U.S. RedCap data showed that COVID-19 affects PWH that are predominantly of older age (mean 51.4 years), African-American (47.5%), and having high rates of comorbidity (80%), with 57.3% of hospitalization, 16.5% of ICU admission, and 9.4 % of mortality.^13^ Data captured from the ISARIC WHO CCP study showed that after adjusting for age, PWH have 47% higher mortality rates (adjusted hazard ratio 1.47, and 95% confidence interval [CI] 1.04-2.25) in England.^14^ The astonishingly rapid spread of COVID-19 spurred the desperate need of COVID-19 vaccines for PWH.

MVC-COV1901 is a subunit vaccine based on the stabilized prefusion SARS-CoV-2 spike protein S-2P (15 mcg) with furin cleavage site mutation and T4 fibritin for trimerization, and formulated in adjuvant composed of TLR9 agonist CpG 1018 and aluminium hydroxide.^15-17^ Previous phase 1 and 2 clinical trials have shown that the MVC-COV1901 vaccine was well tolerated and elicited robust immune responses.^18,19^ In this study, PWH and the matched HIV-negative subjects were compared for the safety and immunogenicity of MVC-COV1901.

## Materials and Methods

### Study design and participants

This is a substudy within a Phase 2, prospective, double-blinded and multi-centre study to evaluate the SARS-CoV-2 vaccine MVC-COV1901. PWH were on stable antiretroviral therapy with CD4^+^ T cell count greater than 350 cells/mm^3^ and HIV viral load less than 10^3^ copies/ml. We compared PWH from a per-protocol immunogenicity subset of the main study with HIV-negative participants of the main study. Participant’s ages ranged from 20-87 and all received two standard doses of 15 mcg MVC-COV1901, administered 28 days apart via IM injection.

Immediate adverse events (AEs), both solicited local and systemic, and unsolicited AEs and adverse events of special interest (AESIs) were recorded throughout the study period.

Immunogenicity was assessed by measuring GMTs and SCRs of neutralizing antibody. The detection and characterization of neutralizing antibodies were performed by central laboratories using validated pseudovirus and/or live virus neutralization assays.^19^ To measure neutralizing antibody titers, wildtype SARS-CoV-2, Taiwan CDC strain number 4 (hCoV-19/ Taiwan/4/2020; Global Initiative on Sharing All Influenza Data accession ID EPI_ISL_411927), was titrated to calculate the 50% tissue culture infective dose (TCID_50_). Vero E6 cells were seeded in 96-well plates (at 1.2×10□ cells per well) and incubated. Starting from a 1:8 dilution, the serum samples were diluted two-fold eight times to a final dilution of 1:1024. Diluted serum samples were then mixed with an equal volume of 100 TCID_50_ per 50 μL of virus. After incubating the serum-virus mixture at 37°C for 1 h, it was added to the wells containing Vero E6 cells. The cells were then incubated at 37°C in a 5% CO_2_ incubator for 4–5 days. The neutralizing titer (NT_50_) was defined as the reciprocal of the highest dilution capable of inhibiting 50% of the cytopathic effect. The NT_50_ results were derived from quadruplicates and calculated with the Reed-Muench method. Neutralizing antibody titers were converted to the WHO Standardized Unit, IU/mL. The conversion is based on the WHO validated NIBSC reference panel.

### Statistical analysis

For the statistical analyses, descriptive statistics were first obtained and used to present sociodemographic and other characteristics. SCRs and 95% confidence interval (CI) were computed for individuals with at least fourfold increase from the baseline. Fisher’s exact test was used to compare seroconversion between the HIV group and the main group. GMTs were estimated from neutralizing antibody titers measured at 28 days after the second dose of the study intervention. GMT ratios, computed as the ratio between the GMT of neutralizing antibodies in the HIV subgroup versus control group, were also estimated. To assess the magnitude of the difference in immune response between the two groups, an analysis of covariance (ANCOVA) model was used. The model included the log-transformed antibody titers at Day 57 as the dependent variable, group (HIV subgroup and main group) as an explanatory variable and adjusted for age, BMI, gender and comorbidity profile. The 95% CI for the adjusted neutralizing antibody titers of each vaccine group was obtained. Then adjusted GMT was back-transformed to the original scale. Adjusted GMT ratio of the two groups and corresponding 95% CI were also estimated. Correlation between GMT and CD4/CD8 ratio, and GMT and absolute CD4^+^ T cell count were analyzed using Spearman’s test. One-way ANOVA was applied for calculating the correlation between GMT and HIV classification stages. Lastly, using age and gender as covariates and a digit-based greedy and nearest neighbor approach with a 1:5 ratio, propensity score matching was employed for robust comparison between 2 groups.

## Results

### Study Population

In the main study, a total of 3854 participants were randomised to treatment groups. Among them, 58 PWH were randomized to the MVC-COV1901 group (Figure 1). One PWH was excluded from the analytic sample due to lack of information on an outcome indicator (i.e. neutralizing antibody titer). There were 882 participants who were HIV-negative and aged 20 to 87 in a per-protocol immunogenicity subset from the main study.

**Figure 1.**
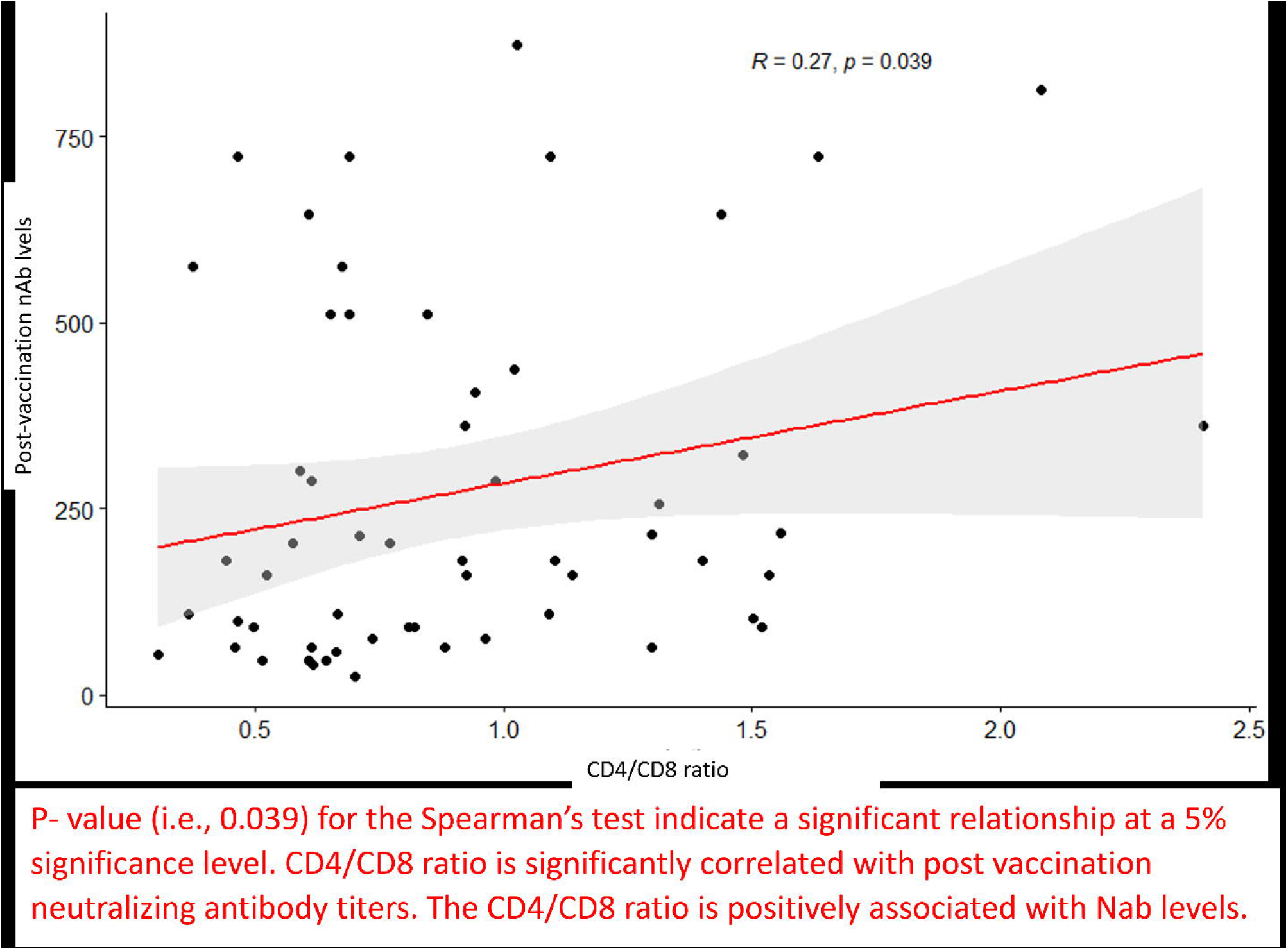
Study design and the trial profile.

The demographic characteristics of the 57 PWH and the 882 comparators from the main study are summarized in Table 1. The mean age of the HIV group and the main group were 38.6 (Interquartile range [IQR] 27-46) and 45.7 (IQR 31-63) years, respectively (p=0.0026); 94.7% and 57.3% were male, respectively (p<0.0001); 19.3% and 11.5% had BMI >= 30 kg/m2, respectively (p=0.087), and 10.5% and 19.4% had comorbidities at baseline, respectively (p=0.116).

**Table 1.** Demographic characteristics of the participants

| Item\MVC lot | MVC-COV1901<br>(HIV positive) | MVC-COV1901<br>Main Study | p-value |
| --- | --- | --- | --- |
| **□ Age (years)** |  |  |  |
| N (Missing) | 57 (0) | 882 (0) |  |
| Mean (SD) | 38.6 (13.1) | 45.7 (16.65) |  |
| Median (IQR) | 36 (19.0) | 43.0 (32.0) | 0.0026 |
| Q1~Q3 | 27.0~46.0 | 31.0 ~ 63.0 |  |
| Min~Max | 23.0~72.0 | 20.0~ 87.0 |  |
| **□ Gender** |  |  |  |
| N (Missing) | 57 (0) | 882 (0) |  |
| Male | 54 (94.7%) | 505 (57.3) | <0.0001 |
| Female | 3 (5.3%) | 377 (42.7) |  |
| **□ BMI (kg/m<sup>2</sup>)** |  |  |  |
| N (Missing) | 57 (0) | 882 (0) |  |
| Mean (SD) | 25.3 (4.9) | 24.9 (4.1) |  |
| Median (IQR) | 24.45 (6.2) | 24.41(5.25) | 0.858 |
| Q1~Q3 | 21.9~28.04 | 22.04~27.3 |  |
| Min~Max | 18.7~39.2 | 14.4~45.2 |  |
| **□ BMI group** |  |  |  |
| N (Missing) | 57 (0) | 882 (0) |  |
| <30 kg/m<sup>2</sup> | 46 (80.7%) | 781 (88.6) | 0.076 |
| >=30 kg/m<sup>2</sup> | 11 (19.3%) | 101 (11.5) |  |
| **□ Comorbidity Category** |  |  |  |
| N (Missing) | 57 (0) | 882 (0) |  |
| Yes | 6 (10.5%) | 171 (19.4) | 0.116 |
| No | 51 (89.5%) | 711 (80.6) |  |

(1) Abbreviations: N=number of subjects in PPI population; SD=standard deviation; Q1=first quartile (25th percentile); Q3=third quartile (75th percentile); IQR=interquartile range; BMI=Body Mass Index; HIV=Human Immunodeficiency Virus.

### Outcomes

#### Safety

Overall, the percentages of participants in each category (solicited adverse events, unsolicited adverse events, and other adverse events after vaccination) were comparable between the HIV group and the main group in all age groups (Table 2-1 & 2-2). The percentages of participants that reported solicited local adverse events were 58.6%, and 72.3% for PWH and main group, respectively. For solicited systemic adverse events, 63.8%, and 53.8%, for PWH and main group. In both PWH and HIV-negative participants, pain or tenderness at injection site was the most common reported local reaction (67.2% and 71.2%, respectively). This event was slightly less common among PWH than HIV-negative individuals. Among systemic reactions, malaise and headache were the most common reaction in both groups but were predominantly mild to moderate in intensity.

**Table 2-1.** Solicited adverse events after any dosing.

|  | All, n (%) |  | ≥ 20 to < 65 years, n (%) |  | ≥ 65 years, n (%) |  |
| --- | --- | --- | --- | --- | --- | --- |
|  | MVC-COV190<br>1<br>(N = 3295) | HIV<br>(N = 58<br>) | MVC-COV190<br>1<br>(N = 2575) | HIV<br>(N = 54<br>) | MVC-COV190<br>1<br>(N = 720) | HIV<br>(N = 4<br>) |
| **Any Solicited Local AEs** | **2381 (72.3)** | **39 (58.6)** | **2030 (78.8)** | **37 (59.3)** | **351 (48.8)** | **2(50.0)** |
| **Pain/Tenderness** | **2346 (71.2)** | **39(67.2)** | **2009 (78.0)** | **37(68.5)** | **337 (46.8)** | **2 (50.0)** |
| Grade 1 | 2237 (67.9) | 37 (63.8) | 1907 (74.1) | 35 (64.8) | 330 (45.8) | 2 (50.0) |
| Grade 2 | 97 (2.9) | 2 (34.4) | 91 (3.5) | 2 (3.7) | 6 (0.8) | 0 |
| Grade 3 | 12 (0.4) | 0 | 11 (0.4) | 0 | 1 (0.1) | 0 |
| **Induration/Swellin**<br>**g** | **347 (10.5)** | **1(1.7)** | **286 (11.1)** | **1(1.9)** | **61 (8.5)** | **0** |
| Grade 1 | 303 (9.2) | 1(1.7) | 246 (9.6) | 1(1.9) | 57 (7.9) | 0 |
| Grade 2 | 42 (1.3) | 0 | 38 (1.5) | 0 | 4 (0.6) | 0 |
| Grade 3 | 2 (0.1) | 0 | 2 (0.1) | 0 | 0 | 0 |
| **Erythema/Redness** | **161 (4.9)** | **0** | **138 (5.4)** | **0** | **23 (3.2)** | **0** |
| Grade 1 | 155 (4.7) | 0 | 132 (5.1) | 0 | 23 (3.2) | 0 |
| Grade 2 | 6 (0.2) | 0 | 6 (0.2) | 0 | 0 | 0 |
| **Any Solicited Systemic AEs** | **1774 (53.8)** | **37 (63.8)** | **1484 (57.6)** | **35 (65.8)** | **290 (40.3)** | **2 (50.0)** |
| **Malaise/Fatigue** | **1186 (36.0)** | **25 (43.1)** | **1036 (40.2)** | **24 (44.4)** | **150 (20.8)** | **1 (25.0)** |
| Grade 1 | 961 (29.2) | 22 (43.1) | 831 (32.3) | 24 (44.4) | 130 (18.1) | 1 (25.0) |
| Grade 2 | 203 (6.2) | 3 (5.2) | 185 (7.2) | 2 (3.7) | 18 (2.5) | 1 (25.0) |
| 1Grade 3 | 22 (0.7) | 0 | 20 (0.8) | 0 | 2 (0.3) | 0 |
| **Myalgia** | **908 (27.6)** | **10 (16.7)** | **757 (29.4)** | **14 (25.9)** | **151 (21.0)** | **2 (50.0)** |
| Grade 1 | 764 (23.2) | 9 (16.7) | 629 (24.4) | 7 (13.0) | 135 (18.8) | 2 |
|  | MVC-COV190<br>1<br>(N = 3295) | HIV<br>(N = 58<br>) | MVC-COV190<br>1<br>(N = 2575) | HIV<br>(N = 54<br>) | MVC-COV190<br>1<br>(N = 720) | HIV<br>(N = 4<br>) |
|  |  |  |  |  |  | (50.0) |
| Grade 2 | 126 (3.8) | 1 (1.7) | 112 (4.3) | 1 (1.9) | 14 (1.9) | 0 |
| Grade 3 | 18 (0.5) | 0 | 16 (0.6) | 0 | 2 (0.3) | 0 |
| **Headache** | **730 (22.2)** | **13 (22.4)** | **631 (24.5)** | **12 (22.2)** | **99 (13.8)** | **1 (25.0)** |
| Grade 1 | 630 (19.1) | 12 (20.7) | 537 (20.9) | 11 (20.4) | 93 (12.9) | 1 (25.0) |
| Grade 2 | 93 (2.8) | 1 (1.7) | 87 (3.4) | 1 (1.9) | 6 (0.8) | 0 |
| Grade 3 | 7 (0.2) | 0 | 7 (0.3) | 0 | 0 | 0 |
| **Diarrhoea** | **497 (15.1)** | **14 (24.1)** | **422 (16.4)** | **14 (25.9)** | **75 (10.4)** | **0** |
| Grade 1 | 411 (12.5) | 12 (20.7) | 350 (13.6) | 12 (22.2) | 61 (8.5) | 0 |
| Grade 2 | 75 (2.3) | 2 (3.4) | 62 (2.4) | 2 (3.7) | 13 (1.8) | 0 |
| Grade 3 | 11 (0.3) | 0 | 10 (0.4) | 0 | 1 (0.1) | 0 |
| **Nausea/Vomiting** | **254 (7.7)** | **6 (10.3)** | **219 (8.5)** | **6 (11.1)** | **35 (4.9)** | **0** |
| Grade 1 | 226 (6.9) | **6 (10.3)** | 192 (7.5) | **6 (11.1)** | 34 (4.7) | 0 |
| Grade 2 | 26 (0.8) | 0 | 25 (1.0) | 0 | 1 (0.1) | 0 |
| Grade 3 | 2 (0.1) | 0 | 2 (0.1) | 0 | 0 | 0 |
| **Fever** | **23 (0.7)** | 0 | **16 (0.6)** | 0 | **7 (1.0)** | 0 |
| Grade 1 | 14 (0.4) | 0 | 8 (0.3) | 0 | 6 (0.8) | 0 |
| Grade 2 | 6 (0.2) | 0 | 6 (0.2) | 0 | 0 | 0 |
| Grade 3 | 2 (0.1) | 0 | 2 (0.1) | 0 | 0 | 0 |
| Grade 4 | 1 (<0.1) | 0 | 0 | 0 | 1 (<0.1) | 0 |

**Table 2-2.** Summary of unsolicited adverse events and other adverse events.

|  | All, n (%) |  | ≥ 20 to < 65 years, n (%) |  | ≥ 65 years, n (%) |  |
| --- | --- | --- | --- | --- | --- | --- |
|  | MVC-COV19<br>01<br>(N = 3295) | HIV in<br>MVC-COV19<br>01<br>(N=58) | MVC-COV19<br>01<br>(N = 2575) | HIV in<br>MVC-COV19<br>01<br>(N = 54) | MVC-COV19<br>01<br>(N = 720) | HIV in<br>MVC-COV19<br>01<br>(N = 4) |
| Unsolicited AEs | 932 (28.3) | 17 (29.3) | 767 (29.8) | 17(29.6) | 165 (22.9) | 0 |
| Related unsolicited AEs | 406 (12.3) | 8 (13.8) | 340 (13.2) | 396 (13.2) | 66 (9.2) | 0 |
| Unsolicited AEs<br>≥ Grade 3 | 93 (2.8) | 3 (5.2) | 86 (3.3) | 96 (3.2) | 7 (1.0) | 0 |
| Related unsolicited AEs<br>≥ Grade 3 | 21 (0.6) | 0 | 20 (0.8) | 24 (0.8) | 1 (0.1) | 0 |
| SAEs | 18 (0.5) | 1 (1.7) | 16 (0.6) | 17 (0.6) | 2 (0.3) | 0 |
| Related SAEs | 0 | 0 | 0 | 0 | 0 | 0 |
| AESI | 1 (<0.1) | 0 | 0 | 0 | 1 (0.1) | 0 |
| VAED | 0 | 0 | 0 | 0 | 0 | 0 |
| AEs leading to study intervention discontinuation | 2 (0.1) | 0 | 0 | 1 (<0.1) | 2 (0.3) | 0 |
| AEs leading to study withdrawal | 1 (<0.1) | 0 | 0 | 0 | 1 (0.1) | 0 |
| Death | 0 | 0 | 0 | 0 | 0 | 0 |

Abbreviations: AE = adverse event; AESI = adverse events of special interest; CI = confidence interval; N = number of participants in the population; n = number of participants with events; SAE = serious adverse event; VAED = vaccine-associated enhanced disease

#### Immunogenicity

After 2 doses of study intervention, the wild type SARS-CoV-2 neutralizing GMT’s (95% CI) on Day 57 (i.e. 28 days after the 2^nd^ dose) in HIV group and main group were 143.48 IU/mL (95% CI 115.5-178.3) and 439.01 IU/mL (95% CI 419.5-459.5), respectively (Table 3). The GMT ratio (95% CI) of main/HIV was 3.06 (95% CI 2.5-3.8). After adjusting for sex, age, BMI category, and comorbidity profile, the GMT’s (95% CI) against wild type SARS-CoV-2 virus were 136.62 IU/mL (95% CI 114.3-163.3) and 440.41 IU/mL (95% CI 421.3-460.4), respectively, and the adjusted GMT ratio (95% CI) of main/HIV was 3.22 (95% CI 2.6-4.1).

**Table 3.** Comparison of wild type SARS-CoV-2 neutralizing antibody (Nab) in IU/mL

| Item\MVC lot | MVC-COV1901 | MVC-COV1901 |
| --- | --- | --- |
|  | HIV positive (n=57) | Main Study (n=882) |
| Nab titer (GMT) in IU/mL | 143.48 | 439.01 |
| 95% CI of Nab titer | 115.5~178.3 | 419.5~459.5 |
|  | Main/HIV |  |
| Nab titer Ratio | 3.06 |  |
| 95% CI of Nab titer Ratio | 2.5~3.8 |  |
| Adjusted Nab titer (GMT) in IU/mL | 136.62 | 440.41 |
| 95% CI of Adjusted Nab titer | 114.3~163.3 | 421.3~460.4 |
|  | Main/HIV |  |
| Adjusted Nab titer Ratio | 3.22 |  |
| 95% CI of Adjusted Nab titer Ratio | 2.6~4.1 |  |

\* estimated using ANCOVA model where sex, age, BMI category, and comorbidity profile are covariates

The seroconversion rates on Day 57 based on the wild type SARS-CoV-2 GMT were 100% in PWH group, and 99.8% in the main group with only two participants in the group failing to seroconvert.

#### Correlation of GMT and CD4/CD8 ratio

CD4/CD8 ratio was positively associated with GMT (R=0.27, p=0.039) (Figure 2). No correlations were found between GMTs and CD4^+^ T cell count (p=0.3), and GMT and HIV classification stages (p=0.44) (Figure S1 and Table S1).

**Figure 2.**
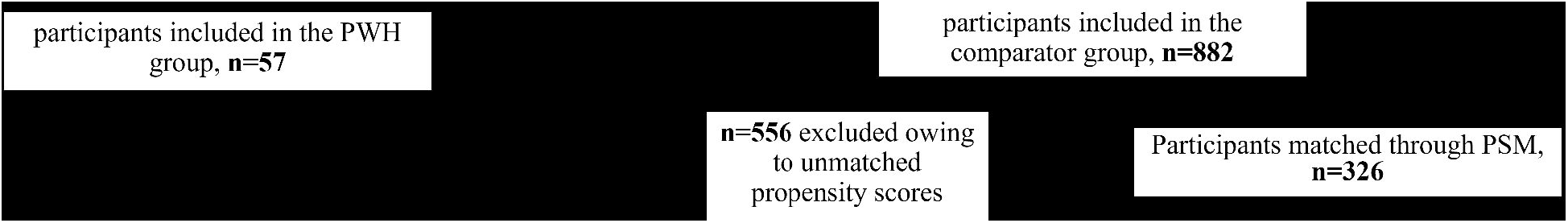
Correlation analysis for CD4/CD8 ratio with post vaccination neutralization antibody titers. P value (i.e. 0.039) for the Spearman’s test indicates a significant relationship at a 5% significance level.

#### Adjusted GMT ratios

Propensity scoring was employed to enable precise ratio determination. A total of 326 subjects in the main study were selected for matching with PWH (Figure S2 and Table S2). The adjusted GMT ratio of matched main/HIV was 3.22 (95% CI 2.6-4.1) (Table S3).

## Discussion

To our knowledge, this is the first study focusing on the safety and immunogenicity of recombinant S-2P protein vaccine against COVID-19 in PWH. The interim analysis demonstrated that in PWH participants aged 20 years and older, two doses of MVC-COV1901 vaccine were safe and well-tolerated, but less immunogenic than in HIV-negative controls. By the time the report was written, all participants having been followed-up for up to six months after the second booster dose.

Previous research raised the possibility that the immune status of PWH negatively modulates the immune responses to COVID-19 vaccines. Specifically, PWH have diminished or less durable responses to hepatitis B and yellow fever vaccination^20-22^ and people with low CD4 cell counts have diminished antibody titers to pneumococcus, influenza, diphtheria, tetanus, and poliomyelitis.^23-25^ Despite these observations, most trials for COVID-19 vaccines have not addressed the PWH subpopulation by subgroup analysis or comparison of PWH with HIV-negative control groups.^30^ These trials include the mRNA-1273 SARS-CoV-2 vaccine phase 3 clinical trial that recruited 179 PWH,^26^ the mRNA COVID-19 vaccine phase 3trial, BNT162b2, that recruited 121 PWH,^27^ and the Ad26.COV2.S vaccine phase 3 trial where 2.8% of the trial population were PWH.^28,29^ Some trials did address PWH: the nCoV-2 (AZD1222) vaccine studied 54 PWH in the United Kingdom, and 103 PWH in South Africa as part of the large phase 2/3 clinical trials of the AstraZeneca COVID-19 vaccine. ^31,32^ All of the participants were on antiretroviral therapy and had high CD4^+^ T-cell counts (median 694 cells/mm^3^ and 695 cells/mm^3^, respectively), and exhibited the same strength of immune response and safety profile, compared to HIV-negative comparators. Similarly a small non-clinical research effort with BNT162b2 involving 12 PWH and 17 healthy donors had demonstrated comparable immunological response.^33^ Finally, Levy et al have studied 143 PWH who received the Pfizer-BioNTech vaccine.^34^ These PWH had antiretroviral therapy and had CD4^+^ T-cell counts of 700 (95% CI 648-757) cells/mm^3^ and 94% of them had undetectable HIV viral load (< 40 copies/ml). Antibodies in these PWH were not inferior to HIV-negative people (98% vs. 99%). In contrast, Spinelli M. et al compared 100 PWH with 100 persons without HIV and found 2.4-fold greater odds in PWH for a nonresponse to pseudovirus by antisera after vaccination with mRNA vaccines.^35^ In addition, the phase 3 clinical trial of NVX-CoV2373 against B.1.351 in South Africa included 201 PWH and the vaccine efficacy was 60% among HIV-negative group versus 49.4% for PWH.^36^ Weaker neutralization antibody responses to Spike protein were demonstrated in this study, but it remains an open question if vaccine efficacy was actually reduced because cellular immunity was not measured.

CD4^+^ T cells orchestrate the response to acute and chronic viral infections by coordinating the immune system. These cells activate B cells to generate efficient neutralisation antibodies, cytotoxic CD8^+^ T cells to kill infected cells, and multiple cells of the innate immune system, and non-immune cells. Thus, CD4^+^ T cells play a key role for the establishment of long-term cellular and humoral antigen specific immunity, which is the basis of life-long protection for many viral infections and vaccines.^37,38^ In addition, CD4^+^ and CD8^+^ T cells produce interferon-gamma (commonly referred to as a ‘‘type 1’’ immune response) which is believed to be protective for the host.^39^ It is therefore a legitimate concern that the immune response could be impeded in PWH with abnormal T cell counts as measured by depleted memory T cells and inversed CD4^+^/CD8^+^ ratios that may be indicative of the response of exhausted cytotoxic T cells toward HIV, and persistent immune activation and inflammation even during stable ART.^40-42^ Nevertheless, generation of high-affinity neutralizing antibodies was a key endpoint in this vaccine study.

Notably, in the study with MVC-COV1901 presented here, CD4^+^ T cell counts did not significantly correlate with GMT in the vaccinated PWH while increasing CD4/CD8 ratios did correlate. The adjuvant of MCV-COV1901, CpG 1018 (a toll-like receptor 9 agonist), may explain this correlation because it binds to the DNA receptor on plasmacytoid dendritic cells, and enhances immunogenicity by stimulating CD4^+^ helper and CD8^+^ cytotoxic T cells.^43,44^ Consistent with these observations, an independent HBV vaccine study in PWH demonstrated that CD4/CD8 ratios > 0.4 were associated with a high rate (86%) of HBV seroconversion.^45^

Despite these indications that cellular immunity may be important for durable protective immunity, there are arguments that long-lived plasma cells may have the potential to produce antibodies for decades in the absence of re-encounter with antigen or specific T cells.^46^ Add to this the 100% of seroconversion in PWH with MVC-COV1901, and it is clear that the role of T cell memory in durable protective immunity against SARS-CoV-2 deserves further study.

Despite the insights generated by our study, some limitations to interpretation may exist. First, the sample size in the PWH group was relatively small. Furthermore, all PWH were on stable ART and had CD4^+^ T cell counts greater than 350 cells/mm^3^ and HIV viral load less than 1000 copies/ml. Thus, extrapolation to people with HIV with lower CD4 counts, or without suppressed HIV viral loads is not suggested. Second, our study was initiated when SARS-CoV-2 was not endemic in Taiwan, and the low viral transmission rate made it difficult to ascertain the efficacy of the vaccine as an exploratory endpoint. Specifically, low levels of 1% of neutralizing antibody titer were detected both at baseline and on Day 57 in the placebo group, suggesting that COVID-19 was rare and natural infection had not boosted the neutralizing antibody titers. ^19^ Third, the short duration of the follow-up period in this study did not allow for assessing the durability of immune responses after Day 57. Fourth, although Th1-skewed immune responses had been demonstrated in the phase I MVC-COV1901 study,^18^ the T-cell responses to the vaccine among PWH were not assessed in this study. Finally, neutralization activities for other SARS-COV2 strains such as Alpha, Beta, Delta, Gamma, and Omicron variants, were not tested and cross-reactivity against emerging Variant of Concerns remains unknown.

In conclusion, this report describes a good safety profile but weaker immunogenicity of MCV-COV1901 in PWH, especially in those PWH with low CD4/CD8 ratios. MCV-COV1901 has emergency authorization use in Taiwan as of July 2021,^19^ and has since advanced to larger clinical trials, including a trial initiated by WHO.^47^ Additional information accumulates from these trials, but further studies are needed to see if PWH require distinct immunization strategies with improved immunogenicity, such as boosters or additional doses,^48,49^ heterologous revaccination,^50^ higher doses as with hepatitis B,^51^ or even a PWH-specific platform for booster shots similar to that for influenza vaccine^52^.

## Data Availability

Data sharing is not applicable to this Article, as it is an interim analysis of data from an ongoing study.

## Contributors

S-HC, CEL and W-SL conceived and designed the study. W-SL, C-YC, W-DL, C-HL, W-CK, Y-HC, H-TC, H-JH, C-TH, N-CW, M-CL, and Y-LL acquired and interpreted the data. CEL, I-CT, and JAGE analyzed the data. S-HC, W-SL, and CEL drafted and prepared the manuscript. CEL, and I-CT had full access to and verified all the data in the study and take responsibility for the integrity of the data and the accuracy of the data analysis. All authors reviewed and approved the final version of the manuscript. S-HC, CEL, W-SL, and T-YL had final responsibility for the decision to submit for publication.

## Acknowledgments

The study was funded by Medigen Vaccine Biologics (study sponsor), the Taiwan Centers for Disease Control and Ministry of Health and Welfare. The sponsor co-designed the trial and coordinated interactions with contract Clinical Research Organization (CRO) staff and regulatory authorities. The CRO operated trial operation to meet the standards of the International Council for Harmonisation of Technical Requirements for Pharmaceuticals for Human Use and good clinical practice guidelines. The IDMC oversaw the safety data and communicated recommendations to the sponsor. The interim analysis was done by the CRO. We thank Stanley Chang, and Charles Chen at Medigen Vaccine Biologics; the investigational staff at National Taiwan University Hospital, Taiwan Taipei Veterans General Hospital, Tri-Service General Hospital, Taipei Veterans General Hospital, Taipei Medical University Hospital, Taipei Municipal Wan Fang Hospital, Linkou Chang Gung Medical Hospital, Taoyuan General Hospital Ministry of Health and Welfare, China Medical University Hospital, Changhua Christian Hospital, National Cheng Kung University Hospital, and Kaohsiung Medical University Chung-Ho Memorial Hospital, for their dedication to this trial; the Clinipace Clinical Research team (Taipei, Taiwan), for their involvement in conducting the trial; Barney S Graham at the Vaccine Research Centre, US National Institute of Allergy and Infectious Diseases, for the development of S-2P pre-fusion protein; Robert Janssen at Dynavax Technologies for providing the CpG 1018 adjuvant and related important intellectual content; members at Protech Pharmaservices (Taipei, Taiwan) for conducting the spike-specific IgG ELISA assay; members at the Department of Laboratory Medicine, Linkou Chang Gung Memorial Hospital (Taoyuan, Taiwan), and members at Institute of Biomedical Sciences, Academia Sinica (Taipei, Taiwan) for conducting the neutralisation assay.

