## supplementary information for "Safety and Immunogenicity of SARS-CoV-2 S-2P Protein Vaccine MVC-COV1901 in People Living with HIV"

Supplementary material

1. Figure S1. Correlation analysis for CD4+ T cell counts with post vaccination neutralization antibody titers.
2. Figure S2. Propensity scores for people who are living with HIV, comparing unmatched participants from main study, and comparing controls with matched propensity score.
3. Table S1. Correlation analysis for HIV classification stage with post vaccination neutralization antibody titers.
4. Table S2. Demographic characteristics for people who are living with HIV and controls with matched propensity scores.
5. Table S3. Comparison of wild type SARS-CoV-2 neutralizing antibody geometric mean titers between people who are living with HIV and controls with matched propensity scores.

Fig S1. Correlation analysis for CD4+ T cell counts with post vaccination neutralization antibody titers.

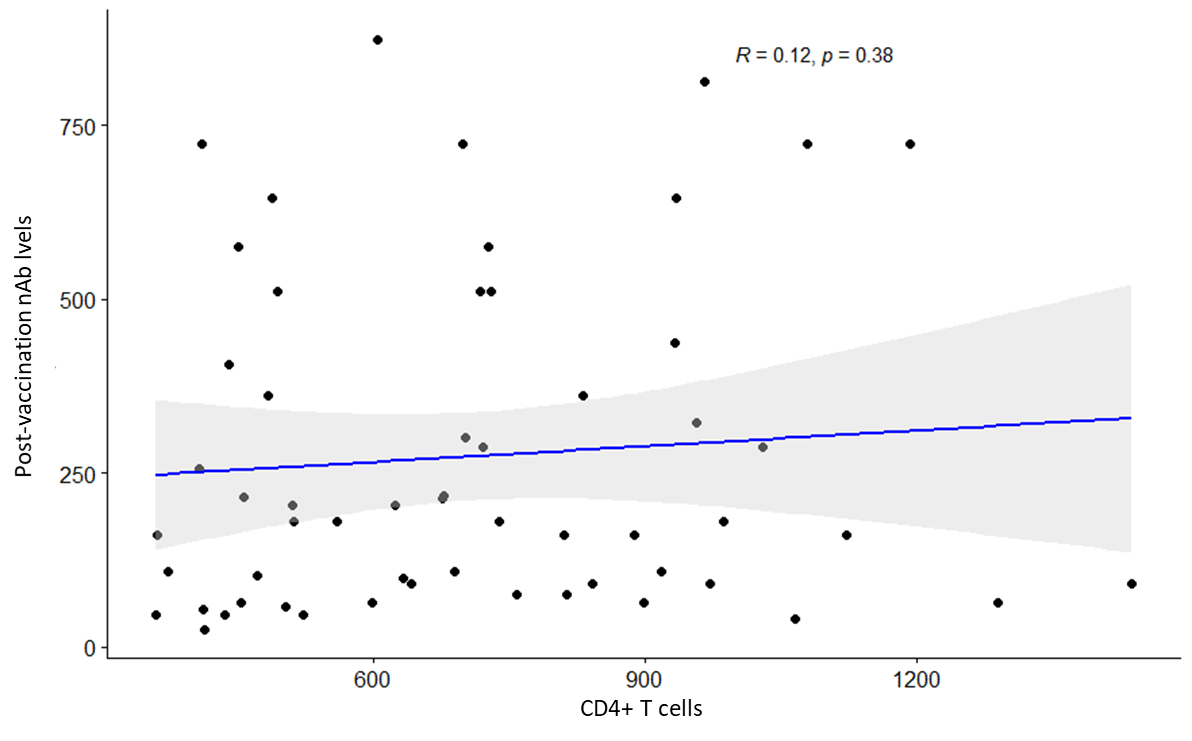

Figure S2. Propensity scores for people who are living with HIV, comparing unmatched participants from main study, and comparing controls with matched propensity score.

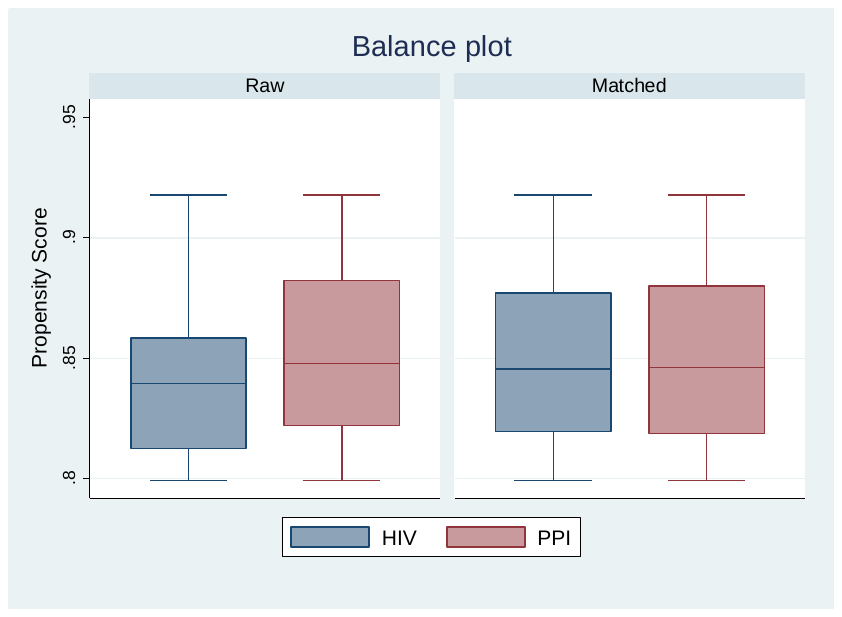

Table S1. Correlation analysis for HIV classification stage with post vaccination neutralization antibody titers.

Correlation between CD4 T cell counts and Post vaccination Nab

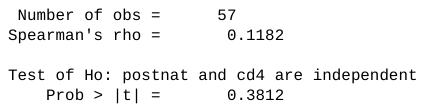

Comparison of CD4 T cell counts (350-500 vs >500)

| **Item** | **CD4 T cell counts** | | | | **p-value** |
| --- | --- | --- | --- | --- | --- |
|  | **350-500 (n=16)** | | **>500 (n=41)** | |  |
|  | **Median** | **IQR** | **Median** | **IQR** |  |
| Post-vaccination Nab | 188.27 | 399.92 | 181.02 | 271.53 | 0.696 |

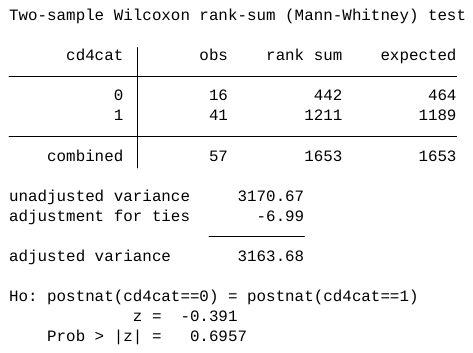

Table S2. Demographic characteristics for people who are living with HIV and controls with matched propensity scores.

| **Item\MVC lot** | **MVC-COV1901** | **MVC-COV1901** | **p-value** |
| --- | --- | --- | --- |
|  | **(HIV positive)** | **Main Study** |  |
| **‧Age (years)** | | |  |
| N (Missing) | 57 (0) | 326 (0) | 0.058 |
| Mean (SD) | 38.6 (13.1) | 42.8 (14.9) |  |
| Median (IQR) | 36 (19.0) | 41 (23.0) |  |
| Q1~Q3 | 27.0~46.0 | 30.0~53.0 |  |
| Min~Max | 23.0~72.0 | 23.0~72.0 |  |
| **‧Gender** |  |  |  |
| N (Missing) | 57 (0) | 326 (0) | 1.000 |
| Male | 54 (94.7%) | 310 (95.1) |  |
| Female | 3 (5.3%) | 16 (4.9) |  |
| **‧BMI (kg/m^2^)** |  |  |  |
| N (Missing) | 57 (0) | 326(0) | 0.167 |
| Mean (SD) | 25.3 (4.9) | 25.9(4.02) |  |
| Median (IQR) | 24.5 (6.2) | 25.4 (5.12) |  |
| Q1~Q3 | 21.9~28.04 | 23.2~28.3 |  |
| Min~Max | 18.7~39.2 | 16.6~40.5 |  |
| **‧BMI group** |  |  |  |
| N (Missing) | 57 (0) | 326 (0) | 0.413 |
| <30 kg/m^2^ | 46 (80.7%) | 277 (84.97) |  |
| >=30 kg/m^2^ | 11 (19.3%) | 49 (15.03) |  |
| **‧Comorbidity Category** |  |  |  |
| N (Missing) | 57 (0) | 326(0) | 0.160 |
| Yes | 6 (10.5%) | 59 (18.1) |  |
| No | 51 (89.5%) | 267 (81.9) |  |
| *(1) Abbreviations: N=number of subjects in PPI population; SD=standard deviation; Q1=first quartile (25th percentile); Q3=third quartile (75th percentile); IQR=interquartile range; BMI=Body Mass Index; HIV=Human Immunodeficiency Virus.* | | | |

Table S3. Comparison of wild type SARS-CoV-2 neutralizing antibody geometric mean titers between people who are living with HIV and controls with matched propensity scores.

| **Item\MVC lot** | **MVC-COV1901** | **MVC-COV1901** |
| --- | --- | --- |
|  | **HIV positive (n=57)** | **Main Study (n=326)** |
| Nab titer (GMT) in IU/mL | 143.48 | 442.97 |
| 95% CI of Nab titer | 115.4~178.3 | 412.5~475.7 |
|  | Main/HIV |  |
| Nab titer Ratio | 3.09 |  |
| 95% CI of Nab titer Ratio | 2.5~3.9 |  |
| Adjusted Nab titer (GMT) in IU/mL | 138.53 | 445.70 |
| 95% CI of Adjusted Nab titer | 116.2~165.1 | 414.3~479.5 |
|  | Main/HIV |  |
| Adjusted Nab titer Ratio | 3.22 |  |
| 95% CI of Adjusted Nab titer Ratio | 2.6~4.1 |  |
| * estimated using ANCOVA model where sex, age, BMI category, and comorbidity profile are covariates | | |

Abbreviations: CI=confidence interval; GMT: geometric mean titers;HIV=Human Immunodeficiency Virus.
